# Are quality medicines affordable? Evidence from a large survey of medicine price and quality in Indonesia

**DOI:** 10.1101/2024.02.21.24303126

**Authors:** Vinky Maria, William Nathanial Tjandrawijaya, Ayu Rahmawati, Yusi Anggriani, Prih Sarnianto, Elizabeth Pisani

## Abstract

**Background:** Since Indonesia implemented one of the world’s largest single-payer health insurance schemes in 2014, the price of many common medicines has fallen dramatically. Public narratives have questioned the quality of low-cost medicines, including those provided free to insured patients. We investigate the relationship between medicine price and quality, and the affordability of medicines paid for out of pocket.

**Methods:** We bought over 1,000 samples of five common prescription medicines -- allopurinol, amlodipine, amoxicillin, cefixime and dexamethasone -- online and from randomly-selected pharmacies and health facilities in four regions across Indonesia, recording price paid, and testing samples for quality using high performance liquid chromatography. We compared prices with the median price for the same medicine; tested for correlation between quality and price, and calculated affordability compared with the district minimum wage.

**Results:** Medicines available in the public procurement system were less likely to fail quality testing than other brands/varieties (4.2% vs 8.3%) but the difference was not statistically significant (p=0.086). There was no other relationship between quality and price, or branded status. Branded generic medicines sold at a large variety of price points, from 0.1 to 23.1 times the median price for the medicine and dose (interquartile range: 0.9 – 4.8, median 1.4).

Unbranded generics traded in a narrower range (range: 0.1 – 3.2; IQR: 0.5 - 1, median 0.8). Medicines were most expensive in the region with the lowest wages, but even there, medicines selling at the 25th centile of available prices cost a maximum of 0.7% of one day’s wage for a course.

**Conclusion:** Though medicine price vary very widely in Indonesia, we found that affordable varieties of common prescription medicines were widely available across the country, and these medicines were no more likely to fail quality testing than those costing several times as much.

## INTRODUCTION

In Sustainable Development Goal 3.8, the nations of the world aspire to ensure “access to safe, effective, quality and affordable essential medicines and vaccines for all”.^1^ In fact, many nations are still far from meeting that goal. According to the United Nations, the proportion of households spending over 10% of their budget on health is increasing, affecting one billion people in 2019.^1^ World Health Organization medicine pricing policy guidelines recommend a number of policies intended to increase the affordability of medicines, including promoting the use of unbranded generic medicines.^2^ While global discussions about fair pricing for medicines are careful to consider the cost of quality assurance,^3,4^ recent, well-publicised cases of cost-cutting in production facilities for lower-priced medicines have raised concerns about the quality of these medicines, feeding in to a narrative, long promoted by manufacturers of more expensive branded medicines, that equates affordable prices with questionable quality.^5,6^

### The Indonesian health system

The relationship between medicine pricing and quality has been a topic of active debate in Indonesia, the world’s fourth most populous nation, and home to one of the world’s largest single-payer health insurance schemes. The mandatory scheme -- Jaminan Kesehatan Nasional or JKN -- was introduced in 2014. At the end of 2023, it reported that over 95% of Indonesians are registered as JKN participants.^7^

Registered users are entitled to free care at all levels for most health conditions. All medicines on the national formulary should also be provided to insured patients for free. Employers or individuals pay a monthly premium; the state covers premia for the poor.

Between 2014 and 2022, most medicines for JKN patients were procured through an online platform known as e-Catalogue. The Ministry of Health (MoH) estimated demand volumes and set a ceiling price for each medicine on the procurement list. With exceptions for products with three or fewer suppliers, any company with a valid market authorisation for an unbranded generic version of listed product could bid at or below that price, provided they undertook to supply demand (at least up to the MoH’s estimate). The lowest bidder in each province won the contract to supply the medicine to that province for one (later two) years; winning prices were published online.^8–10^ As a result of JKN implementation, the price of most commonly-used medicines fell dramatically. This was true not just for unbranded medicines sold through the public procurement system, but also for off-patent medicines marketed by domestic manufacturers using brand names (which we refer to as branded generics).^11^

The Ministry of Health and parliamentarians continued to focus their attention on a sub-set of medicines that remained relatively expensive, contributing to a national narrative about medicines being over-priced in Indonesia.^12,13^ Meanwhile, many people in the pharmaceutical industry reported that the ceiling prices imposed by the government for publicly procured medicines were irrationally low. When the cost of active ingredients, labour or other inputs rose, slim margins were erroded. Manufactuers were reluctant to supply at a loss, leading to stock-outs.^14–17^

In some ways, those stockouts worked in favour of healthcare providers. Before JKN, most patients would pay health facilities out of pocket for any medicines received, contributing to profitability (and in rarer cases sustainability) of service provision. Now, medicines supplied to public primary health centres by the district health bureau are paid for out of general taxation. In other settings, JKN reimburses dispensing health facilities for medicines for chronic conditions, paying the e-catalogue price plus a small margin for handling. All other medicines are expected to be covered by the hopsital or other health provider from claims based on a patient’s diagnosis.^18^ Health facilities are thus incentivised to encourage patients to pay out of pocket for medicines or brands not covered by JKN.^17,19^ Although they are entitled to free medicines, many JKN participants continue to pay out of pocket for medicines, because covered medicines are not available; because they have a preference for other brands; or because they prefer to pay than to access often inconvenient and bureaucratic free services.^20–23^

Previous research in East Java found that Indonesian patients paid widely varying prices for different branded and unbranded versions of cardiovascular and anti-diabetic medicines. All samples passed quality tests, thus, no relationship was found between their price and their pharmacopeial quality.^24^ The authors of that study, which collected 204 samples, suggested that pharmaceutical companies may be protecting investment in quality assurance for very low priced unbranded generic products by selling a branded version of the same product at a higher price.

In this study, we bought over 1,000 samples of five common prescription medicines from physical and online pharmacies and health facilities in four regions across Indonesia. We investigated the affordability of medicine paid for out of pocket; pricing strategies among companies producing multiple similar products; and the relationship between medicine price and quality.

## METHODS

This study uses data collected as part of STARmeds, an investigation of medicine quality in Indonesia, combined with open-access medicine registration data published by the Indonesian national medicine regulator, Badan Pengawasan Obat dan Makanan (BPOM), and statistical data published by Statistics Indonesia.^25,26^

### Study Settings and Data Collection

Detailed methods for the STARmeds study, describing the selection of medicines and sampling areas, construction of the sample frame, and fieldwork and medicines testing, along with all of the data collection forms and laboratory protocols, are published elsewhere.^27^

Briefly, we purposively chose seven sampling areas to reflect Indonesia’s geographic, economic and demographic diversity. These were the Greater Jakarta region, and a large city and a more remote rural district in each of Western, Central and Eastern Indonesia (respectively Medan/Labuhan Batu; Surabaya/ Malang; Kupang/ Timor Tengah Selatan). Within each area, we listed, verified and randomly selected outlets: pharmacies; over-the-counter medicine shops; hospitals; health centers; private doctors, midwives and online stores)

We sampled five medicines – allopurinol, amlodipine, amoxicillin, cefixime and dexamethasone – in tablet or capsule form as available. We also collected dry syrup amoxicillin. We aimed to buy medicines at a variety of price points; each retail outlet was visited by two different mystery shoppers, each targeting a broadly cheaper or more expensive medicine. They signalled their desired price point using phrases such as “Do you have anything a bit more affordable?” or “Is this the best brand you have?”. At healthcare facilities, we sampled overtly, buying all available versions of the target medicine. We also sampled from all registered apps offering instant delivery of medicines using geo-positioning, as well as from online stores of registered physical pharmacies. [The STARmeds study also conducted purposive sampling of medicines sold by unauthorised sellers online. These samples (n=186) were difficult to find; they are not considered in this paper because they do not reflect products easily accessible to most patients.]

Data related to price and product details were entered into a form pre-loaded onto the shoppers mobile phones using the open-source KoboCollect software.^28^

### Quality Testing

For budgetary reasons, we excluded some samples from testing, using a pre-determined triage system (see study archive). For all tested samples, we identified the active ingredient and performed assays (quantifying the active ingredient as a percentage of the labelled dose). All tablets or capsules were tested for dissolution (the percent of labelled active ingredient dissolving in a given time interval). For medicines with doses below 25 mg, (amlodipine and dexamethasone) we also tested for uniformity of content of tablets in the same sample. A sample is a single active ingredient, brand, formulation and dose collected at a single time and place. PT Equilab International, an ISO/IEC 17025: 2017-certified private laboratory in Jakarta, performed the testing between April and November 2022, with real-time supervision from the STARmeds chemist.

Samples were tested using USP reference standards, in accordance with the USP 43 NF 38 monographs. There was no USP monograph for cefixime capsules, so we followed Farmakope Indonesia VI’s adoption of Chinese Pharmacopeia standards.^29,30^ Assay testing was done by high-performance liquid chromatography (HPLC -UV Waters, Aliance 2695 with UV Detector 2489), using columns as specified by USP, while dissolution was by Spectrophotometer-UV/VIS (Shimadzu UV-1800) with the exception of amoxicillin tablets and dexamethasone, where dissolution was tested by HPLC.

A sample was considered out of specification if it failed any quality test it was subjected to. The field data form and the laboratory data were merged on barcode using Stata 18.0 software. Stata 18.0 was also used for reproducible cleaning, coding and to generate descriptive statistics and graphs. Code is provided in the STARmeds repository.

In analyses involving quality, we included 1,088 samples for which we had laboratory data. Because quality of the same brand is unlikely to vary by dose, these include 57 samples of doses that were not specifically targeted in our sampling frame (for example, in quality analyses we included 22 samples of amlodipine 10mg, as well as the 191 samples of the targeted 5mg dose of amlodipine).

### Verification

We provided all 79 market authorisation holders with high resolution photos of every sample of their medicines, asking them to verify that batch numbers, expiry dates and maximum retail prices printed on the primary packaging accorded with their production records. A sample was considered falsified if the market authorisation holder confirmed that the information on the primary packaging did not match their records.

### Price Variation

In order to report comparable measures across medicines with different base price points, we created a standardised ratio. When comparing different brands/manufactuers of the same medicine, we calculated the ratio of the individual sample’s price per smallest counting unit (single tablet or capsule for all medicines except amoxicillin dry syrup, which was priced in 5ml units) to the median price of all samples of the same medicine (active ingredient, dose and formulation). We priced medicines provided free through the public health system at the public procurement price plus an allowed 28% handling margin. When comparing prices charged by different outlets for the identical product, (the same market authorisation holder, brand, dose, formulation), we calculated the ratio of the individual sample to the lowest price paid for that specific product.

In analyses of price variation, we include only the seven combinations of active ingredients specifically targeted in data collection, whether or not we tested them for quality; we excluded non-targeted doses because small numbers did not give a fair picture of price variation across that product.

### Affordability

We adapted methodology developed by WHO/HAI to estimate medicine affordability.^31^ We compared the cost of each medicine with the minimum wage in the district of sampling as follows:

- Considering that low-income patients are generally able to ask for and obtain cheaper brands, we set the price at the 25th percentile of all prices paid for that active ingredient, formulation and dose in the specific sampling area.
- We calculated the number of units needed for a recommended course of treatment of each medicine per month, using defined daily doses recommended by the World Health Organization, and duration of treatment recommended by the Indonesian National Medicines Formulary 2021 for the most commonly indicated condition for each dose and formulation;
- We multiplied the unit cost of each medicine by the estimated number of units needed for a full course of treatment for an acute condition, or a 30.4 day’s supply of medicine for chronic diseases.

Medicines were considered affordable if the required course cost one day’s wage or less. Because minimum wages are set monthly, covering holidays and days off, we calculated one day’s wages as the monthly minimum wage divided by 30.4.

We also compared the cost of a course of treatment with the cost of a month’s supply of other commonly-consumed commodities: the staple food (rice), a common convenience product (instant noodles), and a non-essential item (cigarettes). We used the national average per capita expenditure on these items from data reported in the 2022 round of Indonesia Social and Economic Survey (SUSENAS), a regular survey of over 345,000 households, which is representative at the district level.^32^ Statistics Indonesia calculates per capita expenditure as household expenditure on the commodity divided by the total number of people in the household, regardless of how many people in the household consumed the commodity in question.

### Potential for Cross-subsidization and Quality Effects

To identify the potential for cross-subsidies in product manufacturing or sales, we downloaded BPOM’s database of medicines registered in the Indonesian market, for all versions of the study medicines. We ordered the list by active ingredient, formulation type, market authorization holder and branded status. Any unbranded generic medicine that had a branded equivalent registered to the same market authorization holder was considered potentially cross-subsidised. Other unbranded generics were labelled “unbranded only”.

We compared the prevalence of out-of-specification samples between “unbranded only” generics and unbranded generics that were paired by the market authorisation holder with a brand, testing for the significance of diffrence with a Pearson’s Chi2 test.

### Ethics

This sub-study was registered with the Health Research Ethics Committee, Faculty of Medicine, University of Indonesia (protocol number 20-09-0999). Ethical approvals for the larger STARmeds study are posted in the study repository, where we also describe reporting of suspect products, and other issues with ethical implications.

## RESULTS

### Description of Samples

The distribution of samples tested, by medicine, dose, formulation, geographical area of sampling and source are provided in Supplementary Table 1, Supplementary Table 2 and Supplementary 3. Because we excluded samples differ for different types of analyses, we provide tables for the numbers underlying each figure in our paper. Overall, we included 1,090 samples in analyses of price variations, 1,088 samples in analyses involving quality and 1,003 samples in analyses of affordability.

### Medicine price variations

Medicine prices, including those of unbranded generic medicines, varied very widely, as shown in Figure 1. Across all molecules, unbranded generics sold at a median of 0.8 times the median price for their medicine and dose (range 0.1 – 3.2 IQR 0.5 – 1), and at a median of 3.6 times the public procurement price (range 1.1 – 37.7, IQR 2.4 - 6.5). Branded generics sold at a median of

**Figure 1.**
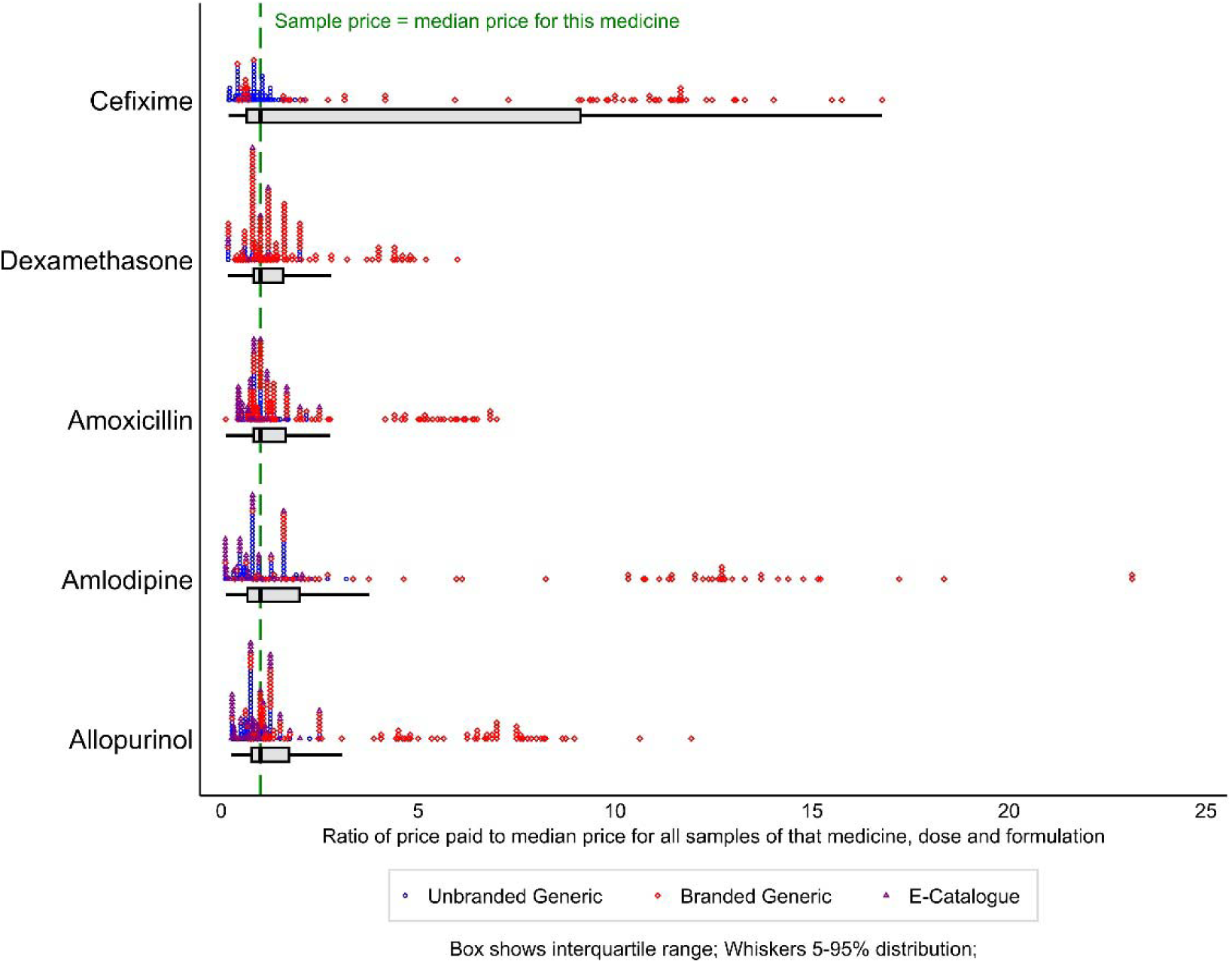
Ratio of sample price to median price for the same medicine, dose and formulation; by active ingredient and branded status.

1.4 times the median price for the medicine and dose (range: 0.1 to 23.1; interquartile range: 0.9 – 4.8), and at a median of 10.6 times the public procurement price (range: 1.3 to 274.5; interquartile range: 4.6– 25.2). The greatest range of both unbranded and branded prices were for amlodipine, an anti-hypertensive medicine often taken daily over the long-term by people at high risk for cardio-vascular disease. Supplementary Figure 1 shows the same data on a log scale, which provides greater clarity of detail at the lower ranges. Supplementary Figure 2 depicts the range of prices across all medicines, by type of outlet.

Although branded medicines were more expensive in general, all study medicines included unbranded generics priced higher than the lowest-priced branded product.

The diversity was not just between brands, but also because different outlets charged different amounts for the identical product. Figure 2 illustrates these difference for the 10 branded and 10 unbranded products (same medicine, dose, formulation and brand) with the widest price variation. Across all products with 5 or more samples, the highest priced seller charged an average of 2.8 times more than the lowest-priced seller for unbranded generics and 1.5 times more for branded products.

**Figure 2.**
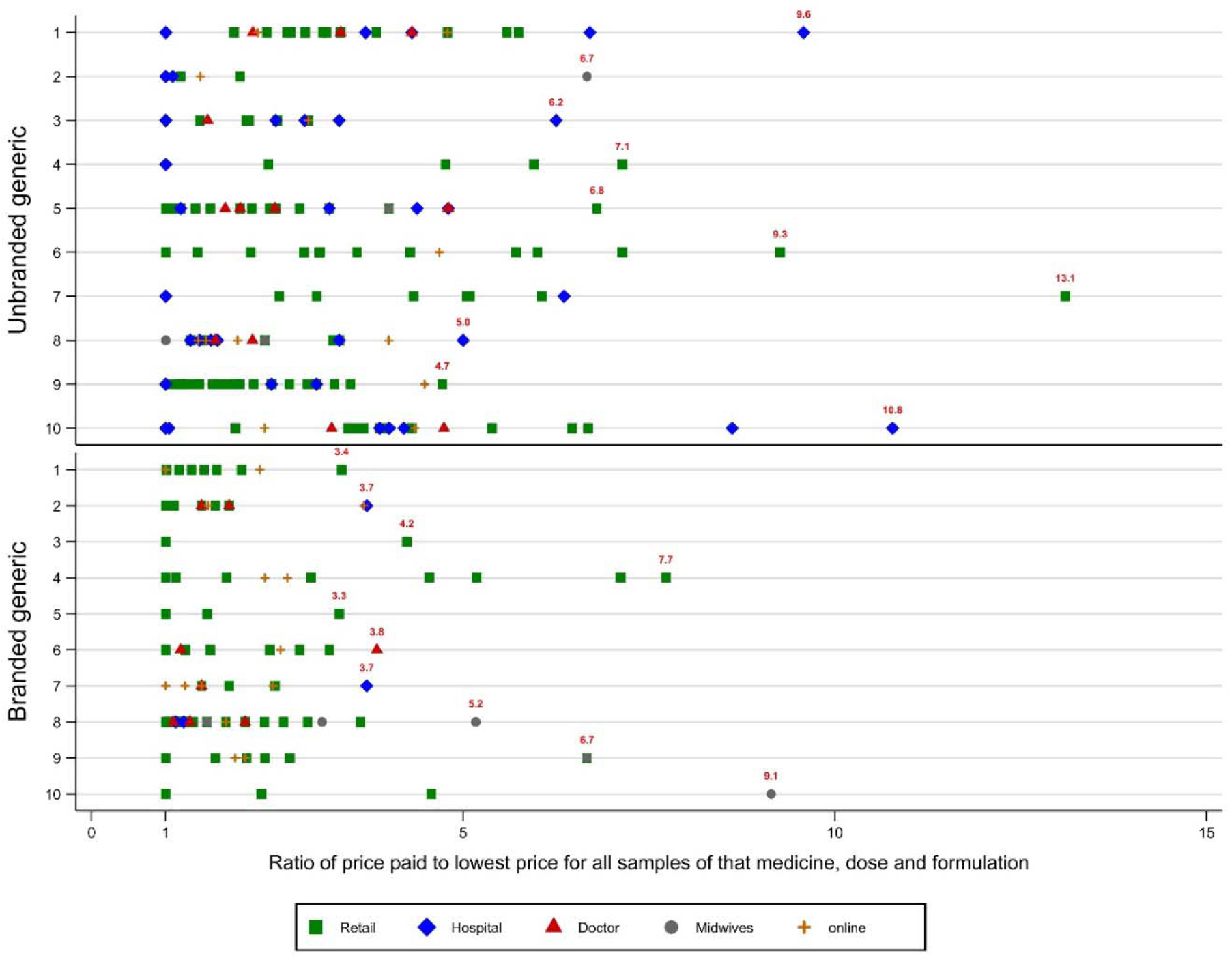
Variation in prices paid for the identical product at different outlets; showing 20 highly variable products.

### Price and Quality

In this analysis, we include 1,088 samples subjected to any chemical testing, sampled from physical outlets or the online stores of registered pharmacies. Of 67 manufacturers, 27 (40.3%) made at least one of the samples that failed a pharmacopeial test in our study, while at least one out-of-specification product was registered to 30 of the 72 market authorization holders (41.7%). Of the 42 market authorization holders who confirmed data of these samples against production records, 3 market authorization reported a total of 6 falsified products, all branded. Full details of quality testing are reported elsewhere, and the sample-level data with all laboratory results are available in the study archive.^33,34^

Here we focus on the relationship between price and quality. Of the 1,088 samples included in the quality analysis, 84 failed any pharmacopeial test, for an unweighted prevalence of 7.7% out-of-specification samples.

Figures 3 plots test outcomes against price (relative to the medicine’s median) for branded and unbranded medicines. In logistic regression, there was no relationship between price and pharmacopeial quality, including after controlling for differences in medicine, district, or source. Samples confirmed as falsified sold at an average of 3.2 times the median price for the medicine, dose and formulation, but there was no difference in price between falsified and non-falsified products of the same brand.

**Figure 3.**
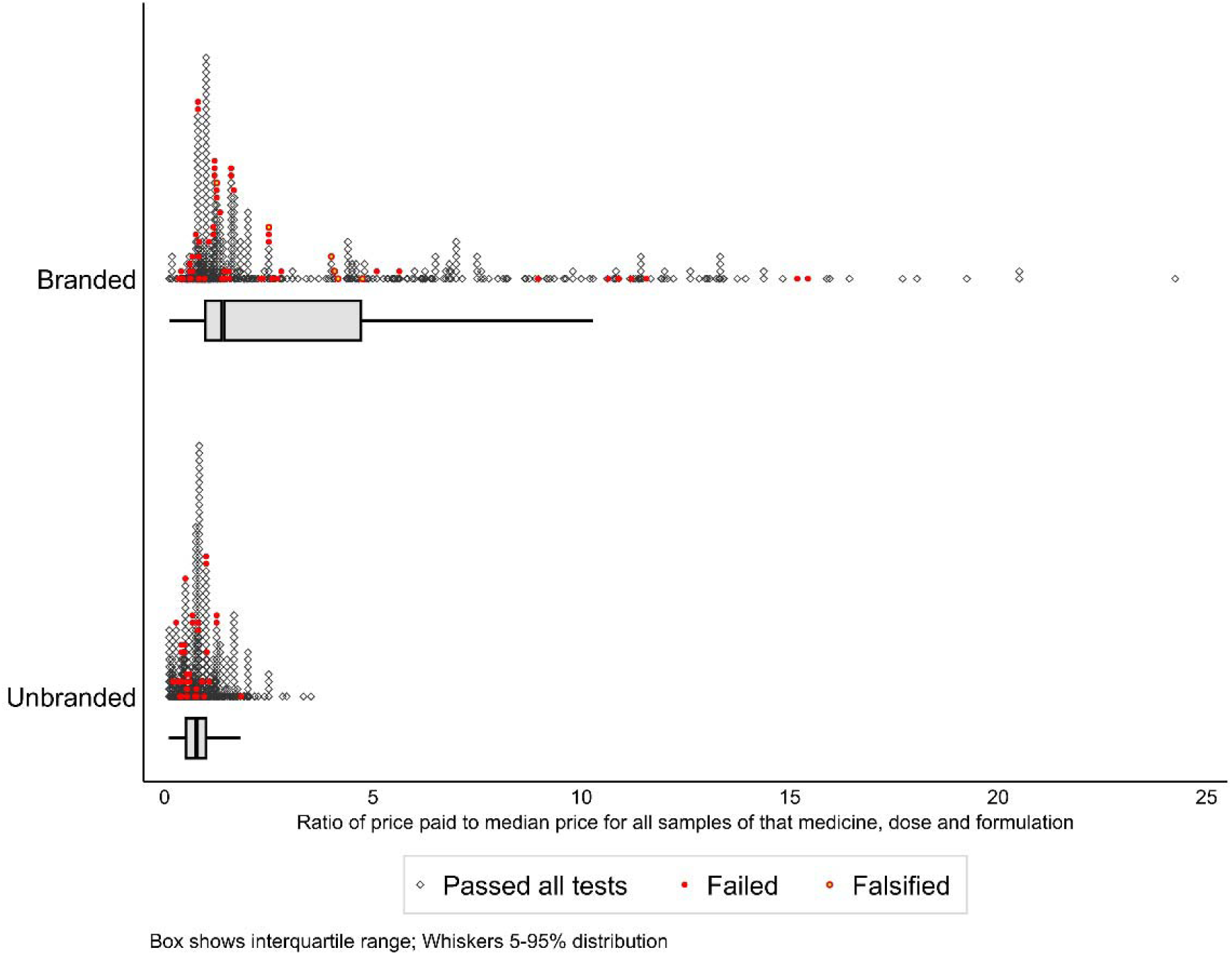
Branded and unbranded samples showing relative price and test results.

There was no significant difference in quality between branded and unbranded products (8.3% vs 7% on pharmacopeial tests alone, p=0.41) including after controlling for differences in medicine, district, or source. However, if we included confirmed falsified samples, branded products were more likely to be categorized as poor quality than unbranded generics (13.3% vs 7.8%, p=0.004). Medicines available free to patients in the public insurance system were less likely to fail testing than medicines paid for out of pocket, but the difference was not statistically significant at the 95% confidence level (4.2% vs 8.3%, p=0.086).

### Affordability of Medicines

A course of medicines priced at the 25th percentile of prices paid locally cost more relative to minimum wages in remote Eastern Indonesia than in other study sites. The outlay was between two and three times higher in remote eastern areas and rural districts than it was in large cities. This was both because medicines were more expensive in East Nusa Tenggara than in other regions, and because the minimum wage was lower in that province. In the district with the highest relative costs, a treatment course of the most expensive medicine in our study cost 2.3 % of the monthly minimum wage, or 0.70 days’ wages (Details are provided in Supplementary Figure 3).

The absolute amount paid for a course of medicines at the 25th percentile of sample prices across all sites ranged from 4,200 rupiah (about USD 0.28 at the 2022 average exchange rate of USD 1 = IDR 14,870) to 34,200 rupiah (USD 2.3). We compared spending on medicines at the 25th percentile with average monthly per capita spending on other commodities reported by Statistics Indonesia. Per capita spending on the staple food, rice, was twice as high as spending on the most expensive course of medicine in the study, and 18 times as much as the cheapest. Spending on instant noodles, a common, low-priced convenience food, was two-thirds of the average course-of-treatment spend for the medicines in our study. Indonesian households spent five times as much per person (including the non-smokers) buying cigarettes, on average, as they did on the average course of treatment in the study (shown in Figure 4).

**Figure 4.**
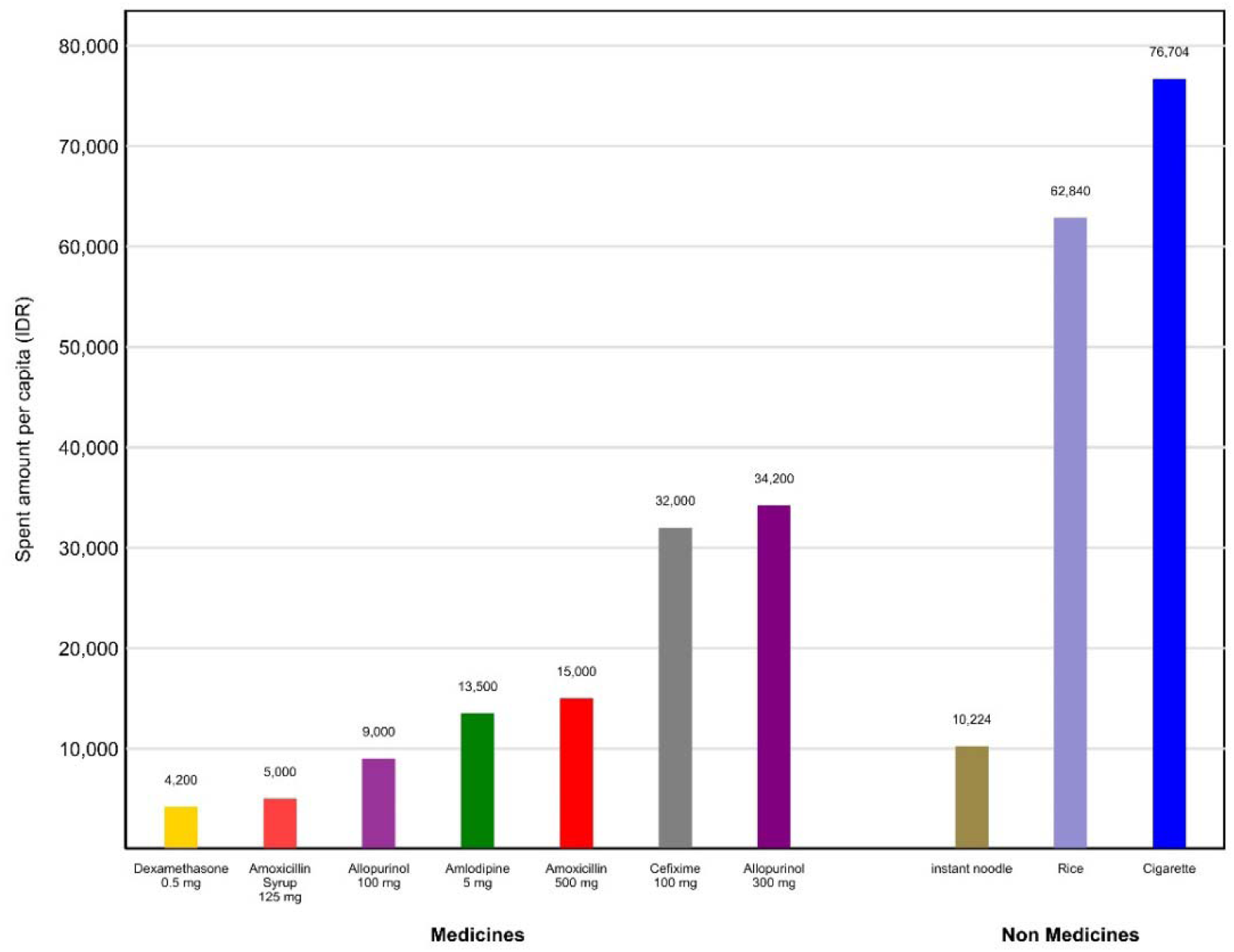
Comparison with montly costs of study medicines and other commodities.

### Potential for cross-subsidization in the Indonesian medicine market

In our study, we collected 246 samples of unbranded generic products whose market authorization holder also registered a branded equivalent (active ingredient and formulation), and 254 samples of unbranded products that had no registered branded equivalent. In order to test the hypothesis that quality of cheaper unbranded products might be protected through cross-subsidisation with more expensive branded products, we compared the quality of these two groups of unbranded generics (see Table 1).

**Table 1.**
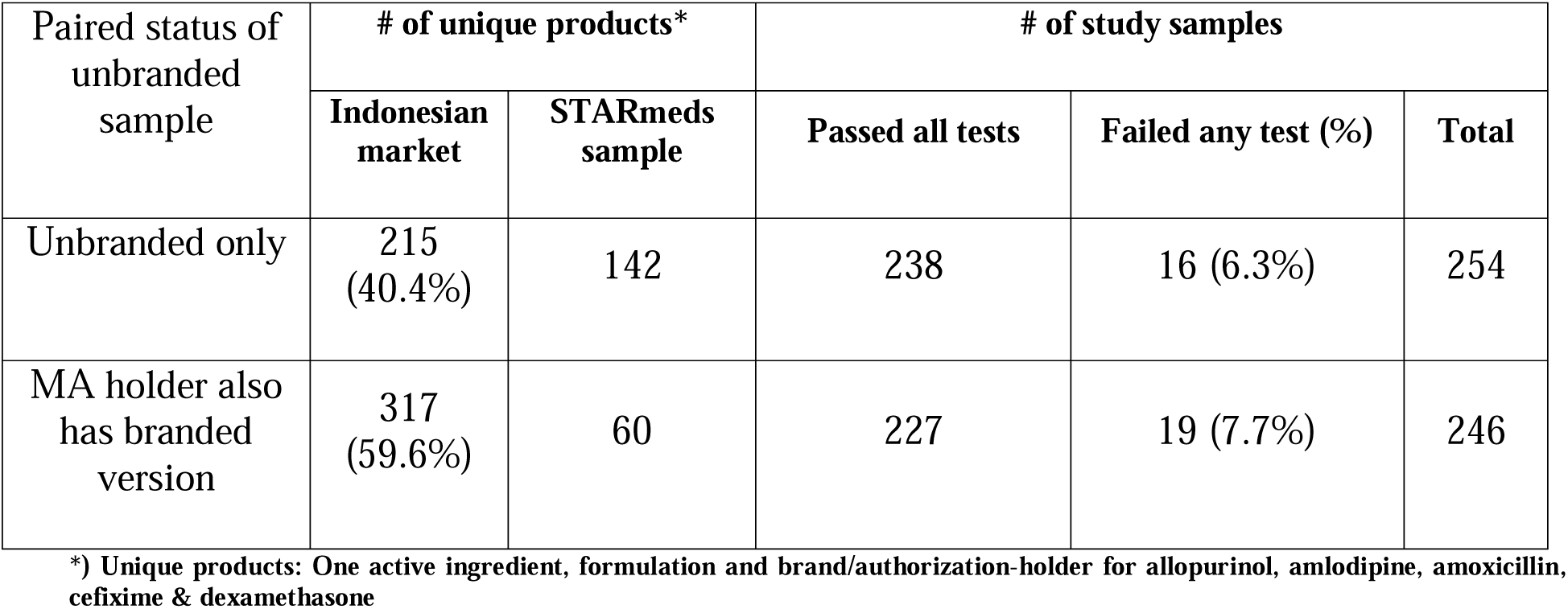
Relationship between cross-subsidy with sample INN and quality testing.

There was no statistically significant difference in testing failure rates between unbranded products that were not potentially cross-subsidised by a branded version and those that were (p = 0.533).

## DISCUSSION

Ensuring access to quality-assured medicines is a core challenge for governments aspiring to provide universal health coverage to citizens, and affordable prices (to the public system and to patients paying out of pocket) are central to securing that access.

Since introducing a mandatory public health insurance scheme in 2014, Indonesia has introduced policies aimed at reducing the price of medicines.^11^ While manufacturers complained prices had become unsustainably low, potentially threatening quality,^35^ politicians continued to focus attention on the obstacle to access represented by higher-priced products.^36^ This led to conflicting public narratives about medicine price and quality, though the perception that publicly-procured medicines are of low quality dominated.^37,38^ The perception was reinforced in 2022, when several low-cost pediatric syrups made in Indonesia, including one provided to insured patients, were found to be contaminated,^39^ allegedly contributing to some 200 child deaths.^40^

In this large study of medicine quality in the Greater Jakarta region and 6 other rural and urban disctricts across Indonesia, we collected information on the actual prices paid by patients paying out of pocket for five common prescription medicines. We also tested samples for quality. We found a great diversity of both branded and unbranded products, selling at a wide array of price points; for all medicines and doses targeted in our study, the most expensive products sold for between five and 21 times the median price, (and between 11 and 173 times the price of their cheapest retail equivalents). Leaving aside outliers and considering just the interquartile range, samples at the 75th percentile sold for multiples of 4.3 times more than those at the 25th percentile. This is a wider range than that identified in studies of price ranges in other countries, including Ireland and China,^41,42^ and is consistent with studies that show that mark-ups on insulin were far higher in Indonesia than in India or Ghana, especially in the private sector.^43^

Britton et al. suggested that the introduction of national health insurance contributed to the segmentation of the Indonesian pharmaceutical market, with multinational companies concentrating their sales on branded medicines, while domestic companies focused on unbranded generics.^14^ However, we found that domestic companies were themselves segmenting the market by selling both branded and unbranded versions of the same medicine at different price points.

Although we identified more out-of-specification medicines than were found in Dewi et al.’s recent study of cardiovascular medicines in East Java, our findings concurred with theirs that there is no evidence of any relationship between price and quality, meaning that patients choosing the cheapest unbranded medicines are not disadvantaged.^24^ Our findings did not, however, support those authors’ speculation that pharmaceutical companies may protect investment in assuring the quality of lower-priced medicines by cross-subsidising through the sale of high-priced brands of the same product. In our study, there was no difference in the quality of unbranded generics that were unique, compared with those that were matched with a more expensive brand.

Accessing free medicines in Indonesia can be time-consuming and inconvenient, so the motivation to pay if quality-assured medicines are affordable is clear. But it is less clear why people are willing to pay such a high premium for some brands when they could choose to buy versions that meet the same pharmacopeial specifications more cheaply. We were not able to interview patients; a major limitation of our study. But we believe at least part of the answer must be that, whatever the facts, Indonesian patients continue to associate low cost with low quality. This phenomenon has been widely reported elsewhere^44^ including in the descriptively titled study “This body does not want free medicines”, an examination of South African patients’ views on unbranded generic medicines procured through state systems.^45^ Our study adds to a body of evidence suggesting that policy-makers should accompany cost-containing procurement policies with strategies to manage consumer perceptions of medicine quality if they wish to increase acceptance of lower-priced medicines.

Similarly, policy-makers should consider how procurement regulations impact on the people and institutions that control patient access to medicines. Again, we did not directly interview healthcare providers, but we note that the highest mark-up on branded medicines were often supplied by hospitals. Previous studies in Indonesia found that physicians, pharmacists and other healthcare professionals influence patients’ choice of medicines, often nudging them towards brands that will maximize revenues from medicine sales, including through incentive payments from pharmaceutical sales teams.^16,24,46^ This is not a universal phenomenon.^47^ However, the dynamic is also at play in the United States and China, which, like Indonesia, have mixed health systems in which the sale of medicines has historically been a profit center.^44,48^

It is concerning that medicine prices were highest in the poorer Eastern regions of Indonesia where incomes are lowest, likely a consequence of high transport costs across the archipelago, as well as of distance from the centres of pharmaceutical production in Java. We note also that patients tended to pay more when buying medicines from doctors and midwives. These health workers are often the principal source of medicines in remote rural areas of Indonesia. However, they are not allowed to buy medicines from distributors. This means they buy at higher retail prices, adding an additional mark-up when selling on to their patients. We believe this restriction should be reconsidered, so that patients in remote areas are not doubly disadvantaged.

Overall, however, we found that Indonesian patients in most settings could access affordable, quality-assured versions of the commonly-prescribed medicines in our study if they chose to. These were available at retail and from health facilities for patients who choose to pay out of pocket, or were obliged to because they were not correctly insured, or because public facilities were out of stock. For every medicine and in every district, a course of treatment could easily be obtained for less than one day’s wages, even by people who cannot use health insurance for administrative or other reasons. For even the most expensive medicine in the study, this is about half as much as households spend per person on another necessary commodity, rice, and just 40% of what the average household spends on cigarettes, a discretionary product.

Indonesia continues to face challenges in delivering health services evenly across the vast, geographically challenging and economically diverse archipelago, with access to hospitals and specialist doctors a particular challenge.^49^ This clearly affects access to medicines that are delivered in tertiary settings.^50^ Further, changes to the medicine procurement system implemented from January 2023, which eliminate consolidated buying through transparent, single-winner tenders, are likely to affect the pharmaceutical market in unpredictable ways. We note also that, in common with virtually all academic studies of medicine quality, we could not afford to test for impurities, meaning that our results may understate the true number of substandard samples. However, our results suggest that at the time of the study, public procurement policies and market dynamics, coupled with relatively strong regulatory oversight of domestic production and the supply chain, were carrying Indonesia closer to the goal of universal health coverage. Indonesians enjoyed access to medicines of good quality at least for these five widely-used medicines (covering four therapeutic categories), whether they choose to queue up for free medicines at public facilities, to buy the cheapest unbranded generic, or to pay over 100 times more for a premium brand.

## Data availability statement

Additional data are available in three locations, all within the STARmeds repository: https://dataverse.harvard.edu/dataverse/STARmeds Supplementary. This includes a replication dataset for **this specific paper** (including the underlying data, the analysis code in Stata format for this paper and supplementary figures).

Data and documentation related to STARmeds fieldwork more generally are also in the study archive. This archive is easiest to use in Tree view. It contains **the sample level data** produced by the STARmeds field study, including raw laboratory data, in csv format. This includes samples collected from illegal online sellers which were excluded from the analysis reported in this paper, using the code <drop if online_wild==1>. Also included are laboratory protocols and a more detailed description of methods. The archive can be accessed at: https://doi.org/10.7910/DVN/RKYICP.

Finally, we provide a **free Toolkit** to help researchers and regulators design and implement medicine quality field surveys using mystery shoppers. The toolkit contains downloadable and adaptable versions of data collection software, field control forms, field worker contracts and other potentially useful documentation. The Toolkit can be downloaded from: https://doi.org/10.7910/DVN/OBIDHJ

## Funding source

The study was funded by UK taxpayers through the UK Department of Health and Social Care and the National Institute for Health Research, under NIHR Global Health Policy and Systems Research Commissioned Awards (https://www.nihr.ac.uk/), grant number NIHR131145. Thank you. The funders had no role in study design, data collection and analysis, decision to publish, or preparation of the manuscript.

Thanks also to United States Pharmacopeia, who provided reference standards at discounted prices.

## Author contributions

VM, YA and EP conceptualised the study. WNT and AR led the analysis, with input from VM and EP. VM and WNT drafted the manuscript. All authors reviewed and commented on the manuscript, and contributed to the final draft.

## Data Availability

Additional data are available in three locations, all within the STARmeds repository: https://dataverse.harvard.edu/dataverse/STARmeds Supplementary. This includes a dataset for this specific paper (including the analysis code in Stata format for this paper and supplementary figures).
Data and documentation related to STARmeds fieldwork more generally are also in the study archive. This archive is easiest to use in Tree view. It contains the sample level data produced by the STARmeds field study, including raw laboratory data, in csv format. This includes samples collected from illegal online sellers which were excluded from the analysis reported in this paper, using the code <drop if online_wild==1>. Also included are laboratory protocols and a more detailed description of methods. The archive can be accessed at: https://doi.org/10.7910/DVN/RKYICP.
Finally, we provide a free Toolkit to help researchers and regulators design and implement medicine quality field surveys using mystery shoppers. The toolkit contains downloadable and adaptable versions of data collection software, field control forms, field worker contracts and other potentially useful documentation. The Toolkit can be downloaded from: https://doi.org/10.7910/DVN/OBIDHJ

https://doi.org/10.7910/DVN/RKYICP

https://doi.org/10.7910/DVN/OBIDHJ

## Acknowledgements

The authors thank Jenny Pontoan, Mawaddati Rahmi, Stanley Saputra and Hesty Utami for contributions to data collection, Esti Mulatsari for overseeing laboratory testing, and Yunita Nugrahani and Sarah Njenga for administrative support. STARmeds colleagues at Imperial College London worked on a parallel study related to the cost of surveillance. We thank Katharina Hauk, Sara Valente de Almeida and Adrian Gheorghe for fruitful collaboration. The Indonesian medicine regulator Badan Pengawas Obat dan Makanan and the national statistics bureau (StatisticsIndonesia, or BPS) actively supported development of our methods. The majority of the data collectors were partners of BPS. We thank them for their hard work.

We also thank members of a multisectoral working group on medicine quality estimation known as PEMO, which groups 12 Indonesian government institutions and 5 professional or industry associations, for advice provided over the course of the study, as well as the members of the STARmeds Study Advisory Group (Michael Deats, Kharisma Nugroho, Yodi Mahendradhata, Raffaella Ravinetto, Selma Siahaan, Val Snewin, Virginia Wiseman and Firman Witoelar) for helpful advice.

