## Supplementary Table 3 for "Are quality medicines affordable? Evidence from a large survey of medicine price and quality in Indonesia"

**Table S3: Number of samples included in analysis of affordability, by location of collection, medicines and dose.**

These analyses (shown in Figure 4 and Supplementary Figure 3). included samples that were not tested for quality. They excluded medicines that were not of doses targeted in the sampling design (because numbers were too small to give a representative spread of price points). They also excluded samples that were provided in the public health system at no cost to patient.

|  | Number of samples | | | | | | | | |
| --- | --- | --- | --- | --- | --- | --- | --- | --- | --- |
|  | Greater Jakarta | North Sumatra | | East Java | | NTT | | Online | **Total** |
| Medicine & dose |  | Rural | Urban | Rural | Urban | Rural | Urban |  |  |
| Allopurinol 100mg | 45 | 12 | 23 | 20 | 29 | 11 | 19 | 31 | **190** |
| Allopurinol 300mg | 22 | 0 | 12 | 2 | 10 | 2 | 4 | 5 | **57** |
| Amlodipine_10mg | 0 | 0 | 0 | 0 | 0 | 0 | 0 | 0 | **0** |
| Amlodipine 5mg | 58 | 12 | 22 | 18 | 28 | 7 | 20 | 25 | **190** |
| Amoxicillin 500mg | 44 | 9 | 24 | 21 | 28 | 10 | 20 | 16 | **172** |
| Amoxicillin, dry syrup | 19 | 2 | 13 | 1 | 10 | 2 | 4 | 2 | **53** |
| Cefixime_100mg | 41 | 8 | 20 | 16 | 22 | 10 | 15 | 18 | **150** |
| Cefixime_200mg | 0 | 0 | 0 | 0 | 0 | 0 | 0 | 0 | **0** |
| Dexamethasone_0.5mg | 43 | 11 | 25 | 19 | 33 | 9 | 17 | 34 | **191** |
| Dexamethasone_0.75mg | 0 | 0 | 0 | 0 | 0 | 0 | 0 | 0 | **0** |
| **Total** | **272** | **54** | **139** | **97** | **160** | **51** | **99** | **131** | **1,003** |

NTT: Nusa Tenggara Timur
