## Supplementary Table 2 for "Are quality medicines affordable? Evidence from a large survey of medicine price and quality in Indonesia"

**Table S2: Number of samples included in analysis of quality by price, by location of collection, medicines and dose.**

This analysis (results shown Figure 3) only included samples that were tested for quality. Because we expect the quality of products to be similar across brands regardless of dose, we included medicines that were not of doses targeted in the sampling design. For samples provided free in the public system, we substituted the ecatalog price, plus the permitted 28% margin as a handling fee.

|  | Number of samples | | | | | | | | |
| --- | --- | --- | --- | --- | --- | --- | --- | --- | --- |
|  | Greater Jakarta | North Sumatra | | East Java | | NTT | | Online | **Total** |
| Medicine & dose |  | Rural | Urban | Rural | Urban | Rural | Urban |  |  |
| Allopurinol 100mg | 43 | 14 | 26 | 20 | 31 | 12 | 19 | 31 | **196** |
| Allopurinol 300mg | 21 | 0 | 12 | 3 | 11 | 2 | 4 | 5 | **58** |
| Amlodipine_10mg | 3 | 0 | 4 | 0 | 4 | 1 | 5 | 5 | **22** |
| Amlodipine 5mg | 55 | 13 | 25 | 18 | 27 | 7 | 21 | 25 | **191** |
| Amoxicillin 500mg | 44 | 11 | 29 | 22 | 28 | 11 | 22 | 16 | **183** |
| Amoxicillin, dry syrup | 22 | 5 | 17 | 2 | 9 | 2 | 8 | 2 | **67** |
| Cefixime_100mg | 38 | 9 | 20 | 15 | 20 | 10 | 15 | 17 | **144** |
| Cefixime_200mg | 7 | 2 | 3 | 1 | 0 | 1 | 2 | 3 | **19** |
| Dexamethasone_0.5mg | 42 | 12 | 27 | 19 | 34 | 10 | 18 | 34 | **196** |
| Dexamethasone_0.75mg | 4 | 1 | 2 | 2 | 0 | 0 | 0 | 3 | **22** |
| **Total** | **279** | **67** | **165** | **102** | **164** | **56** | **114** | **141** | **1,088** |

NTT: Nusa Tenggara Timur
