## Supplementary Table 1 for "Are quality medicines affordable? Evidence from a large survey of medicine price and quality in Indonesia"

**Tables S1: Number of samples included in analyses of price variation.**

This analysis included samples that were not tested for quality. We excluded medicines that were not of doses targeted in the sampling design (because numbers were too small to give a representative spread of price points). For samples provided free in the public system, we substituted the ecatalog price, plus the permitted 28% margin as a handling fee.

**S1a: By location of collection, medicines and dose (Figures 1 and 2 and Supplementary Figure 1)**

|  | Number of samples | | | | | | | | |
| --- | --- | --- | --- | --- | --- | --- | --- | --- | --- |
|  | Greater Jakarta | North Sumatra | | East Java | | NTT | | Online | **Total** |
| Medicine & dose |  | Rural | Urban | Rural | Urban | Rural | Urban |  |  |
| Allopurinol 100mg | 46 | 14 | 28 | 21 | 32 | 12 | 22 | 31 | **206** |
| Allopurinol 300mg | 23 | 0 | 12 | 3 | 11 | 2 | 4 | 5 | **60** |
| Amlodipine_10mg | 0 | 0 | 0 | 0 | 0 | 0 | 0 | 0 | **0** |
| Amlodipine 5mg | 60 | 14 | 26 | 19 | 30 | 8 | 23 | 25 | **205** |
| Amoxicillin 500mg | 46 | 11 | 30 | 22 | 30 | 11 | 24 | 16 | **190** |
| Amoxicillin, dry syrup | 20 | 4 | 17 | 2 | 11 | 2 | 7 | 2 | **65** |
| Cefixime_100mg | 42 | 9 | 21 | 16 | 24 | 10 | 17 | 18 | **157** |
| Cefixime_200mg | 0 | 0 | 0 | 0 | 0 | 0 | 0 | 0 | **0** |
| Dexamethasone_0.5mg | 45 | 12 | 29 | 20 | 36 | 10 | 21 | 34 | **207** |
| Dexamethasone_0.75mg | 0 | 0 | 0 | 0 | 0 | 0 | 0 | 0 | **0** |
| **Total** | **282** | **64** | **163** | **103** | **174** | **55** | **118** | **131** | **1,090** |

NTT: Nusa Tenggara Timur

**S1b: By outlet type, medicines and dose (Figure 2, Supplementary Figure 2)**

| Source | Greater Jakarta | North Sumatra | | East Java | | NTT | | Online | **Total** |
| --- | --- | --- | --- | --- | --- | --- | --- | --- | --- |
|  |  | Rural | Urban | Rural | Urban | Rural | Urban |  |  |
| Pharmacy | 202 | 24 | 128 | 70 | 130 | 37 | 75 | - | **666** |
| OTC medicine shop | 43 | 3 | 1 | - | - | - | - | **-** | **47** |
| Primary health centre | 5 | 6 | 5 | 5 | 2 | 4 | 8 | **-** | **35** |
| Hospital | 22 | 12 | 8 | 14 | 21 | 12 | 19 | **-** | **108** |
| Doctor | 8 | 10 | 14 | 5 | 13 | - | 16 | **-** | **66** |
| Midwife | 2 | 9 | 7 | 9 | 8 | 2 | - | **-** | **37** |
| Online | - | - | - | - | - | - | - | 131 | **131** |
| **Total** | **282** | **64** | **163** | **103** | **174** | **55** | **118** | **131** | **1,090** |

OTC: Over the counter
NTT: Nusa Tenggara Timur
