## Supplementary figures and images for "Are quality medicines affordable? Evidence from a large survey of medicine price and quality in Indonesia"

### Supplementary Figure 1

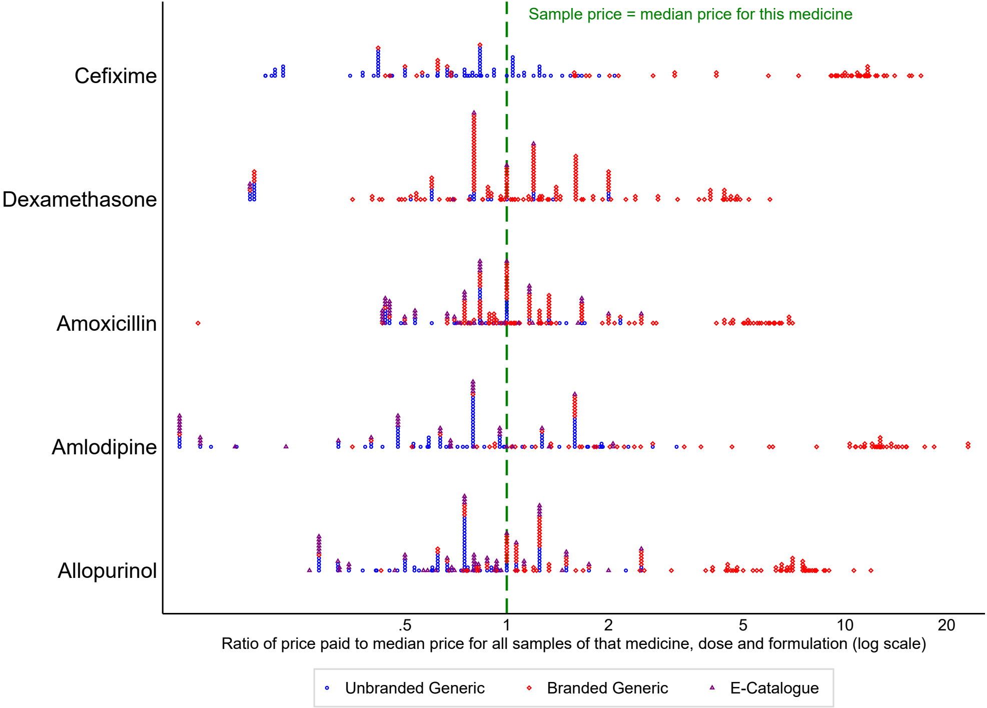

### Supplementary Figure 2

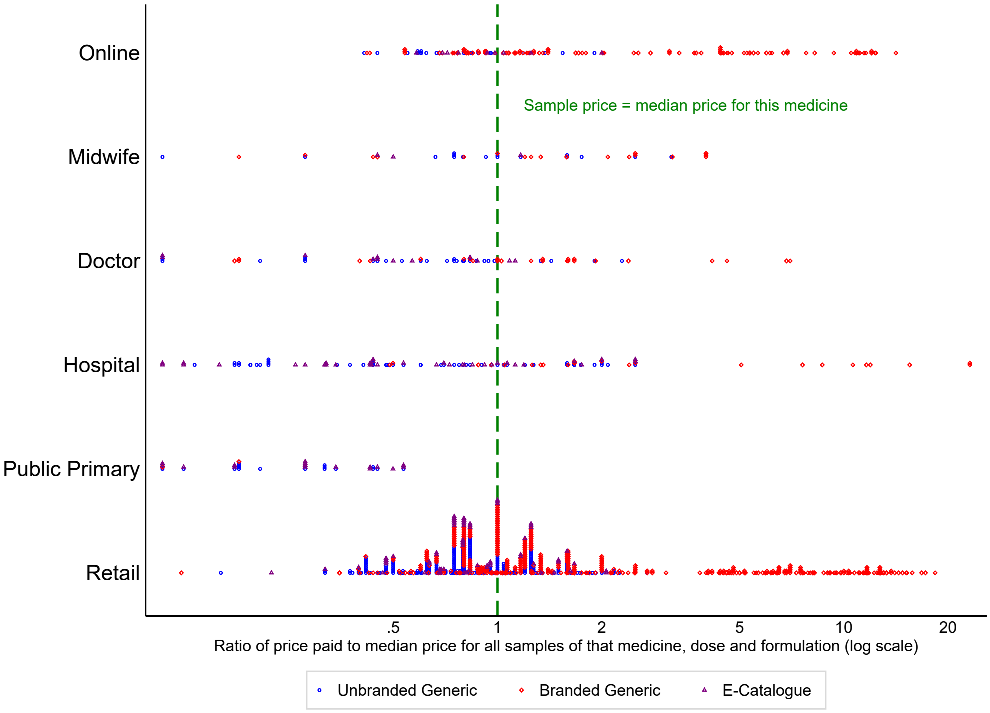

### Supplementary Figure 3

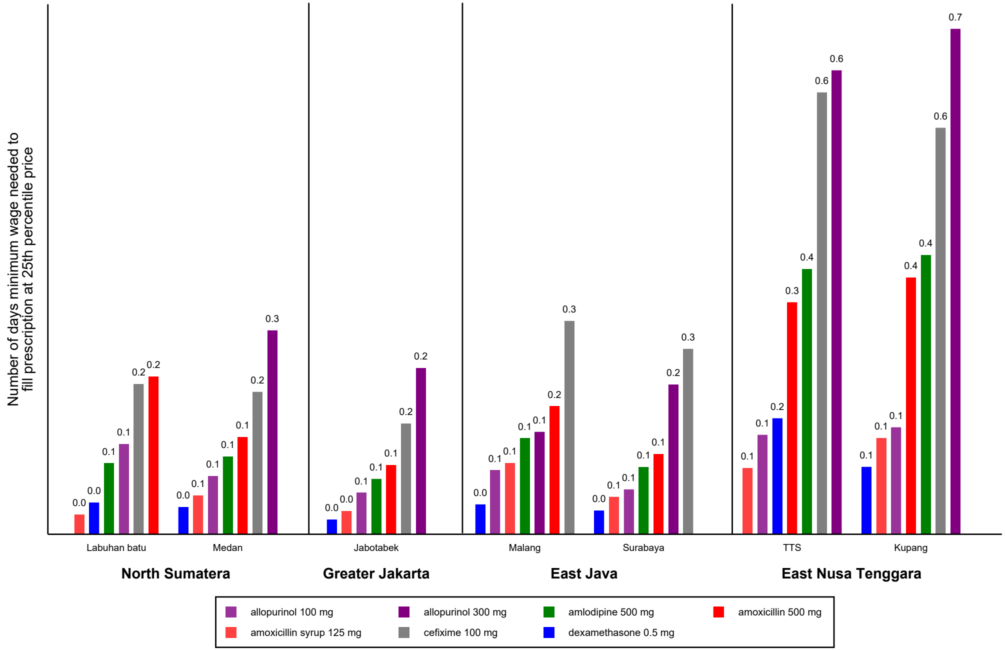
